# Evaluation of Clinical Threshold Policies for Cataract Surgery and the Regional Variation in Rates of Surgery

**DOI:** 10.1101/2021.03.24.21254261

**Authors:** J. Lisle, N. Ansari, M. Jabir

**Affiliations:** Department of Ophthalmology, Rotherham General Hospital, Sheffield; Department of Ophthalmology, Royal Hallamshire Hospital, Sheffield

## Abstract

**Background/Aims:** To characterise the national patterns in rates of cataract surgery across England, review clinical thresholds against NICE guidance, and determine how far variable access to cataract surgery is attributable to regional variation in policy stringency and social deprivation.

**Methods:** 127 Clinical Commissioning Groups (“CCGs”) provided cataract surgery data and threshold policies in response to a Freedom of Information request. Local cataract surgery rates were grouped by threshold stringency and analysed on an age group-corrected basis. ANOVA testing was used to assess effect of policy stringency on regional rates of cataract surgery.

**Results:** In England, rates of cataract surgery vary threefold across CCGs, from 1,980 to 6,427 per 100,000 population over 60, with a standard deviation (784.76) of 22% of the mean value, 3,598. Threshold policies vary across CCGs: 33 had no policy, 45 utilised policies accessible on the basis of Quality of Life (“QoL”) impact, and 39, against NICE guidance, required a Visual Acuity (“VA”) threshold be exceeded. Rates of surgery by CCG were negatively correlated with restrictiveness of policy (η^2^=0.18, p<0.01), and positively correlated with the Index of Multiple Deprivation (IMD), (r^2^□=□0.11, p<0.01). Prior approval processes are not significantly associated with reduced rates of surgery.

**Conclusion:** Over two-thirds of CCGs continue to use threshold-based policies for access to cataract surgery, with one-third doing so solely on the basis of VA requirements, despite NICE guidance to the contrary. For NHS operations, variation in policy restrictiveness accounts for more of the variation in surgery rates than socioeconomic deprivation.

## Introduction

Cataract is the primary cause of blindness globally, and the second leading cause of visual impairment^1^. Surgery is the only effective treatment^2^; and is the most commonly performed surgical procedure in the UK with more than 400,000 cases performed each year^3^. It has excellent outcomes, and is associated with improvements in visual acuity (VA), visual function, and quality of life (QOL)^4–6^.

Unfortunately, rates of cataract surgery across England demonstrate significant geographical variability, suggesting inequitable access and creating an emerging public impression of a “postcode lottery”^7,8^. Although absolute rates of surgery have increased to meet rising demand, variability has also widened over time, with rates of cataract surgery ranging from 328 to 1,166 per 100,000 of total population in 2015/16^9^, compared to the earlier period 1998-2003 when rates ranged more narrowly between local authority areas, from 172 to 548 per 100,000 population^10^. The cause for the discrepancy across regions remains largely unclear. In the US, population studies have linked age, deprivation, race, UV exposure, and urban vs rural residency^11^; but to-date, only social deprivation has been positively correlated with higher rates of cataract surgery in the UK (r^2^□= □0.24 in 2007)^10^.

In England, 135 Clinical Commissioning Groups determine health policy for local populations of typically a few hundred thousand people. The policies include whether and what thresholds are set for access to cataract surgery, and are expected to comply with guidance from the National Institute of Clinical Excellence (“NICE”). A 2012 evaluation of CCG commissioning policies highlighted marked variation of commissioning policies for cataract surgery across CCGs, and the use of arbitrary VA thresholds to determine access^7^. Since then, the Royal College of Ophthalmologists published evidence-based guidance in 2015^12^ recommending access to surgery on the basis of visual function and not visual acuity, and in 2016 NICE guidance was updated to advise “not to restrict access to cataract surgery on the basis of visual acuity”. This was following the results of the NICE systematic review and economic modelling for cataract surgery^13^, which found that immediate first-eye surgery is cost-effective, even with no immediate health-related quality of life gain. Because of the progressive nature of cataracts, for the majority of symptomatic patients it is not cost-effective to delay surgery until a visual acuity threshold is met. Although subsequent reports have highlighted increasingly restrictive commissioning policies^14^, there has been no reevaluation of commissioning patterns since NICE guidance was updated. Furthermore, to-date no study has assessed for any association between the regional variation of CCGs’ thresholds for cataract surgery, and the established variation in rates of surgery.

In this study, we describe CCGs’ commissioning policies for cataract surgery and evaluate their compliance with NICE guidance. We characterise the variation in rates of cataract surgery across England, and explore any association with restrictiveness of policy, to determine whether threshold policies are contributing to unequal access to surgery. Finally, we examine the effect of social deprivation and the impact of prior approval processes, where these are in place.

## Methods

A Freedom of Information (FOI) request was made to 135 CCGs in England in July 2020. Each CCG was asked to provide their clinical threshold policy for cataract surgery, and for the financial year 2019-20, to provide the following:

- The number of prior approval requests the CCG received for cataract surgery;
- The number of prior approval requests for cataract surgery that the CCG approved;
- The CCG’s total number of cataract operations carried out.

Healthcare Resource Groups (HRG) is a clinical coding system used in hospitals across England. Cataract activity data was collected using cataract-related HRG codes, the complete list of which is attached in Appendix 1. Two authors independently reviewed the responses and assessed each CCG’s cataract commissioning policy threshold criteria. Specified lifestyle and visual acuity requirements were noted, as well as thresholds for second eye surgery. Each policy’s date of publication was recorded, along with the nature of any referenced evidence, if present.

Rates of cataract surgery were calculated for each CCG by collating number of cataract operations with the 2019/20 registered population for each CCG. This data is published by NHS Digital online publications^15^ as an age-stratified dataset, allowing us to control for variation in age demographics by expressing operation rates for the over over-60 population of each CCG.

Policies were grouped according to their compliance with current NICE guidance not to restrict access on the basis of visual acuity. Accordingly, three groups of increasing restrictiveness were formed, based on whether a patient could access surgery based on visual acuity, or quality of life impact:

- Those with no policy
- Policies that can be satisfied by a QOL impact (i.e. those with QOL only, or QOL *or* VA requirements)
- NICE – incompliant policies requiring a VA impact (i.e. those with VA only, or VA *and* QOL requirements). Patients in this group with a VA above the requirement would be unable to access surgery, regardless of the severity of the impact on their quality of life.

To assess the impact on restrictiveness of policy on cataract surgery rates, analysis of variance (ANOVA) testing was performed on the cohorts, and a Tukey’s test for post-hoc analysis of pairwise differences between the groups. The Eta-squared measure of effect size was calculated to give an estimate of the proportion of variance accounted for by the variation in policy restrictiveness.

The Index of Multiple Deprivation (IMD) is a calculated measure of social deprivation, incorporating seven distinct domains of deprivation including income, employment, and health. It is published as an anonymised dataset by the Office for National Statistics^16^, with each CCG receiving a score. To determine the effect size of deprivation on rates of cataract surgery, Pearsons’ correlation coefficient was calculated for IMD scores and surgery rates.

## Results

Of the 135 CCGs contacted, 127 (94%) responded. Ten (7%) did not provide information on cataract surgery activity and were excluded, to leave 117 CCGs (87%) for analysis.

### Variation in commissioning policy

There was wide variability of policy criteria between CCGs (Table 1). Thirty-three CCGs (28%) had no policy in place restricting access. Of those with policies in place, 45 utilised NICE-compliant thresholds based on quality of life impact: a small minority, 6, with solely a QOL requirement, and 39 allowing for consideration of visual acuity *or* lifestyle factors. One third of CCGs (39) restricted access to surgery on the basis of visual acuity, against current NICE recommendations. Eight of these did so based on VA alone, and 31 required both a visual acuity *and* QOL threshold to be reached. In CCGs mentioning a VA threshold, Snellen chart VA requirements were either 6/9 or 6/12.

**Table 1:** Rates of cataract surgery and CCG budget allocation for cohorts of stringency of policy

| Policy Type | Number of CCGs | Cataract procedures/<br>100,000 over 60 population | Standard<br>Deviation |
| --- | --- | --- | --- |
| **No Threshold Policy** | **33** | **4,039** | **947.74** |
| **NICE-compliant QOL threshold** | **45** | **3,631** | **641.73** |
| QOL only | 6 | 3,642 |  |
| QOL or VA | 39 | 3,629 |  |
| 6/9 | 18 | 3,767 |  |
| 6/12 | 21 | 3,511 |  |
| **NICE-incompliant VA threshold** | **39** | **3,186** | **549.74** |
| Visual Acuity alone | 8 | 2,870 |  |
| 6/9 | 1 | 2,311 |  |
| 6/12 | 7 | 2,950 |  |
| Visual Acuity and QOL | 31 | 3,268 |  |
| 6/9 | 24 | 3,231 |  |
| 6/12 | 7 | 3,394 |  |
| **Total** | **117** | **3597.86** | **784.76** |

Rates of cataract surgery ranged varied across CCGs, from 1,980 to 6,427 per 100,000 population over 60 years’ age (3.24-fold variation), with a standard deviation (784.76) of 22% of the mean value, 3,598. Rates of surgery were highest in CCGs with no threshold policy (4,039, n=33), followed by those with QOL requirements limiting access (3631, n=45), and finally those restricting on the basis of visual acuity (3185, n=39) (Table 1). There was a statistically significant difference between groups as determined by ANOVA (F(2, 114) = 12.8, p=<0.001). Post-hoc analyses using Tukey’s HSD showed that rates of cataract surgery were significantly higher in CCGs with no policy restricting access, when compared to CCGs with a QOL (p=0.037) or VA (p<0.001) requirement, and that there was a significant difference between the QOL and VA requirement groups (p=0.020). The effect size (η ^2^ = 0.183) demonstrated that variation in policy accounted for 18.3% of the total variance in surgery rates seen. Interestingly, there was no overall difference in surgery rates between those setting the VA threshold at 6/9 and those setting it at 6/12.

Deprivation was significantly associated with increased rates of cataract surgery (R^2^=0.11, p<0.01), accounting for 11% of the standard devation.

**Figure 1:**
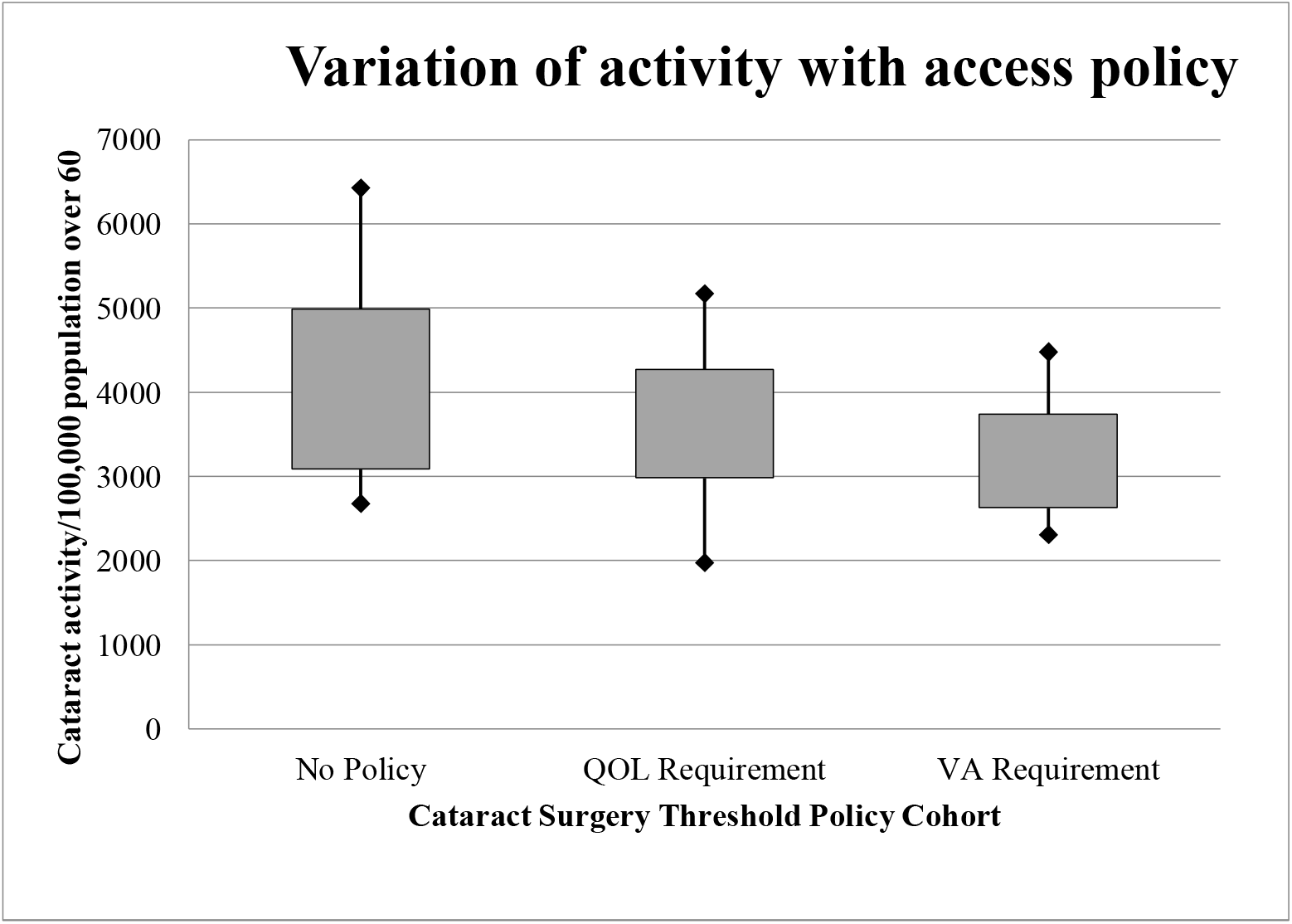
Rates of cataract surgery by policy stringency. The shaded boxes extend to 1 standard deviation above and below the mean, which divides the data set. The whiskers extend to the minimum value and maximum value.

### Quality of Life measures

There was substantial heterogeneity in specified QOL measures across CCGs. Precise wording varied across policies, but QOL considerations could be categorised into general areas (Table 2). Policies were most likely to consider glare and binocular driving, with few considering falls risk or caring needs.

**Table 1:** Number of CCG policies considering different QOL measures

| Policy QOL parameter | Number of CCGs considering |
| --- | --- |
| Binocular driving | 36 |
| Glare | 32 |
| Occupation | 22 |
| Reading | 19 |
| Medical assessment (e.g. medical retina or glaucoma) | 19 |
| Reduced contrast sensitivity | 14 |
| Activities of daily living | 12 |
| Caring needs | 7 |
| Falls | 6 |

### Second Eye Surgery

Only four CCGs made no provision for second eye surgery in their cataract commissioning policies. The majority (64) required the same criteria as for the first eye, with a further eight adding anisometropia in addition to the first eye surgery criteria. Eight CCGs had markedly more restrictive criteria for second eye surgery, with a best-corrected minimum visual acuity requirement of 6/24 in the second eye.

### Questionnair es

Six CCGs utilised assessment questionnaires which numerically scored VA and QOL impact, with a minimum total score threshold. One questionnaire tool, used by four CCGs, was scored such that a minimum VA of 6/9 needed to be reached regardless of QOL measures. Another questionnaire required a minimum score in both VA and QOL domains, and the final questionnaire’s threshold could be reached with QOL impact score alone. Accordingly these CCGs were grouped, irrespective of their use of questionnaires, based on these VA and QOL requirements.

### Prior Approval

Prior approval requests totalled 33,306, of which 30,649 (92%) were approved, representing 7.3% of the total cataract activity (Table 3). Thirteen CCGs reported more than 10% of 2019/20 activity generated from prior approval requests, and six CCGs had all of their cataract activity generated from prior approval processes. These six CCGs had a lower mean rate of cataract surgery per 100,000 over-60 population (M=3429) when compared to the other 111 CCGs (M= 3608), however the difference was not significant (p=0.56).

**Table 3:** Evidential basis of cataract surgery policies

|  | Number of CCGs |  |  |
| --- | --- | --- | --- |
|  | No Evidence | Evidenced, but not current NICE guidance | References current NICE guidance |
| NICE Compliant | 15 | 15 | 15 |
| NICE Non-Compliant | 25 | 8 | 6 |
| Percentage compliant | 37.5% | 65% | 71% |

### Use of evidence

Of the 84 CCGs with cataract commissioning policies, a quarter (21) cited current NICE guidance as supporting evidence. Curiously, six of these are restricting access on the basis of visual acuity (Table 3). Twenty-three policies referenced evidence, but not current NICE guidance, and 40 did not cite any supporting evidence. Given the paucity of evidence for the threshold policies, the majority were surprisingly recently updated: all but seven were published after the 2016 NICE guidance.

## Discussion

### Salient findings

Over two-thirds of CCGs continue to restrict access to cataract surgery through clinical thresholds, with wide variation of policy criteria across CCGs. This study confirms the previously reported geographical variation in rates of cataract surgery, with a three-fold difference between the lowest and highest rates of surgery in the over 60 population and a standard deviation of 22% of the mean value. Although a degree of variability is to be expected due to differences in regional population characteristics, these figures account for age and it is difficult to reconcile them with what might be considered reasonable within the context of an equitable health service. Presence of a threshold policy, increasing restrictiveness of policy, and a higher level of socioeconomic deprivation are all significantly associated with reduced rates of surgery. Differences in deprivation across CCGs account for 11% of the total deviation, with variation in policy accounting for 18.3% of the total variance seen, suggesting that commissioning patterns are contributing more to regional variation in NHS cataract surgery rates than deprivation.

Despite four years having elapsed since the 2016 publication of current NICE guidance, a significant number of CCGs do not reflect NICE guidance in their policy, with one third restricting access on the basis of visual acuity despite NICE guidance to the contrary^13^. There is little or no evidential basis to many of the policies: although 92% of policies were updated after the 2016 NICE guidance, only a quarter of CCGs referenced the publication, with 48% not citing any evidence as the basis for their commissioning policy. Eleven CCGs still consider cataract surgery a “procedure of limited clinical value”, despite its well-established excellent clinical outcomes and cost-effectiveness.

A notable proportion of cataract activity (7.3%) is from prior approval. A 2019 investigation by the BMJ^14^ suggested that this proportion was increasing over time (7.0% in 2016/17 and 10% in 2017/18), but with a notably higher proportion of prior approval activity for 2018/19 (22%), perhaps due to differences in the selection of responding CCGs. We found no significant difference between CCGs using prior approval and those not. With over 90% of cataract activity not through prior approval processes, this study suggests that access is being limited largely by the presence and stringency of threshold policies themselves, rather than the way these are processed.

Heterogeneity of QOL considerations across policies made direct comparison difficult. A 2012 evaluation of threshold policies for cataract surgery^7^ found a similarly substantial variability over the same domains, suggesting that these have not been revisited in almost a decade. The most common factor considered across CCGs is driving, however due to DVLA driving requirements being based on a minimum visual acuity of 6/12^18^, this is essentially a surrogate visual acuity requirement. While this study shows that thresholds do impact access, and the impact is greater if policies are more restrictive, there is evidence to suggest that prioritization tools using QOL measures to assess visual function (as opposed to visual acuity) may be beneficial to allocate resources to patients with greater clinical need^19^, and reduce waiting times^20^. Indeed, it is unclear whether inconsistency in policies may be, in part, due to existing prioritization of competing health needs that vary across CCGS. If the NHS is to introduce prioritization for cataract surgery based on QOL impacts, these requirements need to be clearer and more consistent between CCGs.

### Strengths and Limitations

The FOI request had a good response rate (94%), and association between restrictiveness of policy and rate of surgery is highly significant. The number of CCGs mentioning a visual acuity requirement (78) is consistent with a 2019 BMJ report^14^ (76), and the proportion of CCGs with no policy (28%) concordant with a 2017 RCOphth survey of clinical leads (34%)^17^. The variation described here is notably similar to that found in the 2012/13 NHS Atlas of Variation^8^, which found a 2.9-fold variation of 1596 to 4610 operations per 100,000 population over 65 (M=3033).

The effects of IMD and policy variation found in this study leave much of the variance unexplained, despite correcting for age, and the study has several limitations. We are dependent on the nature and quality of the FOI response data, and cannot exclude variation in the way these data were collected or processed across CCGs. Secondly, responses made no distinction between first-and second-eye surgery. Our analysis is based upon first-eye criteria, and although only 10% of CCGs had more restrictive policies for second-eye surgery, there is the possibility of distinct patterns of regional variation of rates between the two. Thirdly, private cataract surgery is, necessarily, omitted from this study because it bypasses CCG threshold criteria. However, regional patterns of private work may contribute to the variability in rates of surgery across CCGs, and explain the discrepancy between the effect size of deprivation found in this study (11%), and that previously reported by Keenan *et al* (24%)^10^, who used hospital episode statistics inclusive of private activity. Fourthly, the decision to set the level for age-correction at 60 years was based, in part, on a median age of cataract surgery of 67.7 years in the US^11^. Although results were significant regardless of correction for age, the decision for setting for over 60 as opposed to over 55, or 70, is arbitrary. Finally, in our interpretation of these results, we assume that policy is translated into practice, when adherence to these policies by referring ophthalmologists is not necessarily guaranteed. If clinicians were only loosely adhering to policy, this would perhaps explain the lack of effect of more restrictive VA requirements. A 2017 survey of ophthalmologists^17^ found that for the majority (73%), there was no specific monitoring of adherence to thresholds to access. On the other hand, this study found no association between reduced rates of surgery and prior approval processes which circumvent clinician autonomy and drive higher compliance to policy, suggesting that policies are adhered to in practice.

## Conclusion

This study confirms previous work identifying wide variation across CCGs in both rates of cataract surgery, and criteria for access. Furthermore, this is the first study that the authors are aware of reporting a significant association between the presence and restrictiveness of cataract surgery criteria, and the established geographical variability in rates of cataract surgery. It suggests that, against national guidance, commissioners continue to limit access to surgery to many patients with capacity to benefit, using arbitrary thresholds and with almost half of all policies having no evidential basis. A notable proportion of CCGs still utilise resource-intensive prior approval processes for cataracts, or consider surgery a “procedure of limited clinical value”. These findings are contrary to the established clinical benefit and cost-effectiveness of cataract surgery and current NICE guidance, and hard to reconcile with the principle of equity in healthcare.

## Ethics

Ethical approval was not required for this study, as it is based on publicly accessible information relating to patient populations and involved not patient level intervention.

## Data Availability

No additional data available

## Appendix 1

Z30A Complex, Cataract or Lens Procedures, with CC Score 2+

BZ30B Complex, Cataract or Lens Procedures, with CC Score 0-1

BZ31A Very Major, Cataract or Lens Procedures, with CC Score 2+

BZ31B Very Major, Cataract or Lens Procedures, with CC Score 0-1

BZ32A Intermediate, Cataract or Lens Procedures, with CC Score 2+

BZ32B Intermediate, Cataract or Lens Procedures, with CC Score 0-1

BZ33Z Minor, Cataract or Lens Procedures

BZ34A Phacoemulsification Cataract Extraction and Lens Implant, with CC Score 4+

BZ34B Phacoemulsification Cataract Extraction and Lens Implant, with CC Score 2-3

BZ34C Phacoemulsification Cataract Extraction and Lens Implant, with CC Score 0-1

